# (Hydroxy-) Chloroquine, G6PD deficiency, and sex differences in mortality during the COVID-19 pandemic: An ecological study in Peru

**DOI:** 10.1101/2025.10.09.25337648

**Authors:** Marius Krämer

## Abstract

**Background:** Peru implemented a nationwide chloroquine/hydroxychloroquine (CQ/HCQ) treatment policy during the early phase of the COVID-19 pandemic. Whether these drugs trigger hemolytic crises in individuals with glucose-6-phosphate dehydrogenase deficiency (G6PDd), a genetic condition primarily affecting men, with a relatively high prevalence in certain regions, is a matter of debate. This study investigated whether the CQ/HCQ policy contributed to sex-specific differences in mortality, potentially mediated by regional G6PDd prevalence.

**Methods:** I conducted an ecological analysis using both all-cause excess and COVID-19 mortality data at the state level for adults aged ≥45 years. Using sex differences in age standardized mortality rates (ASMR) as the outcome in two-way fixed effects (TWFE) models, interaction analyses were performed to examine the influence of regional G6PDd prevalence, CQ/HCQ availability, and elevated healthcare system strain during pandemic waves with the CQ/HCQ recommendation.

**Results:** Male all-cause excess and COVID-19 mortality consistently exceeded female mortality, with the largest sex differences coinciding with periods with CQ/HCQ treatment recommendations. The TWFE models revealed that states with higher G6PDd prevalence, greater CQ/HCQ availability, and elevated healthcare system strain presented increased male–female mortality gaps. These results are consistent with the hypothesized mechanism in which oxidative stress from both COVID-19 infection and CQ/HCQ exposure contributed to hemolytic crises among (undiagnosed) G6PD-deficient outpatients, potentially misidentified as severe COVID-19 during periods of healthcare strain.

**Conclusion:** While causality cannot be established owing to the ecological design, these findings suggest that pandemic treatment policies may have unintended, sex-specific consequences if population-specific genetic vulnerabilities are overlooked. They underscore the broader public health implication that drugs considered safe under normal conditions may have unanticipated risks during infectious crises that induce additional oxidative stress, emphasizing the importance of integrating genetic risk awareness into future emergency response frameworks.

## Introduction

Mortality during the COVID-19 crisis has differed between males and females across countries and regions, reflecting both biological vulnerabilities and contextual factors [1, 2]. Some studies show higher mortality among men [3–5], whereas others reveal complex patterns, including periods or locations where women experienced equal or higher mortality [6, 7]. Given the biological plausibility of sex-specific drug risks, treatment policy may represent an overlooked driver of these patterns.

At the beginning of the pandemic, chloroquine (CQ) and hydroxychloroquine (HCQ) were seen as potential COVID-19 treatments. However, international trials led by the WHO were discontinued in July 2020 because of safety concerns and a lack of efficacy [8]. While most countries remained cautious, Peru adopted CQ/HCQ widely. Initial protocols restricted use to moderate or severe cases [9], but by mid-April 2020, HCQ was authorized for mild outpatient cases [10]. Peru retained HCQ in official guidelines until September 2020, months after international trials had halted its use [8, 11]. Given that Peru has one of the highest all-cause excess mortality rates globally [12], it provides a unique context for studying the impact of treatment policies.

The safety profile of CQ and especially HCQ in the context of COVID-19 has been debated. In a meta-analysis of 14 trials testing CQ/HCQ in hospitalized patients, it was associated with an 11% (95% CI 2–20%) increase in all-cause mortality [13]. An observational preprint study by the Peruvian Medical Research Institute (IETSI) found that compared with standard care, HCQ combined with azithromycin was associated with an 84% (95% CI 12–202%) increase in all-cause mortality in hospitalized COVID-19 patients [14]. Evidence from outpatient studies generally reported few safety concerns, but these trials consistently excluded individuals with contraindications to CQ/HCQ [15], often explicitly mentioning glucose-6-phosphate dehydrogenase deficiency (G6PDd) as one such exclusion [16–19].

Among the factors that may influence CQ/HCQ-related risk, G6PDd is particularly relevant: an X-linked enzymatic disorder that renders hemizygous males especially vulnerable, whereas females are usually carriers or only partially deficient; G6PDd can predispose individuals to acute hemolytic anemia when exposed to oxidative stress, which may be triggered by infections or certain drugs [20]. While CQ/HCQ use in malaria-endemic regions has not been associated with widespread hemolytic events, according to a systematic review of the available evidence [21], case reports during the COVID-19 pandemic document acute hemolytic anemia in G6PDd patients treated with HCQ [22–26].

Moreover, COVID-19 itself has been reported to be associated with acute autoimmune hemolytic anemia, often presenting with delayed hemolytic symptoms that may be overlooked due to the predominance of respiratory manifestations [27].

The combination of the oxidative stressors COVID-19 and CQ/HCQ may increase the risk of acute hemolytic anemia among G6PDd patients. In particular, during outpatient CQ/HCQ use, hemolysis symptoms may have been delayed or mistaken for COVID-19 symptoms (e.g., dyspnea, fever), complicating detection, especially in an underdeveloped and heavily strained healthcare system. Furthermore, Peruvian treatment protocols with CQ/HCQ did not include warnings about potential adverse reactions or contraindications for G6PDd patients. Consequently, given that G6PDd disproportionately affects males, the nationwide CQ/HCQ policy could plausibly have sex-specific consequences for mortality.

To my knowledge, no prior study has examined whether national COVID-19 treatment policies contributed to sex-specific differences in all-cause excess or COVID-19 mortality. This ecological study addresses this gap by testing two hypotheses:

1. If CQ/HCQ triggers acute hemolytic anemia in G6PDd COVID-19 patients and Peru’s healthcare system failed to manage these reactions, periods of widespread CQ/HCQ use should coincide with increased male‒female mortality differences. Within these periods, increased healthcare system strain is expected to further amplify these differences.
2. If regional G6PDd prevalence modifies risk, areas with higher prevalence should show stronger male–female mortality differences during periods of widespread CQ/HCQ use.

By integrating publicly available all-cause and COVID-19 mortality data with treatment policy information and regional G6PDd prevalence estimates [28], this study explores the plausibility of the described mechanism.

## Data and Methods

For this analysis, I used Python 3.11, primarily Pandas (v2.2.3) for data handling and Statsmodels (v0.14.1) along with Linearmodels (v5.4) for statistical analysis.

### Data

All datasets are publicly available and were aggregated at the state and week levels. Mortality data from 2017–2019 were used to construct baseline mortality, while all other datasets cover 2020–2022. The compiled dataset includes 24 states over 157 iso-weeks.

### Mortality data

Estimating the true mortality burden of COVID-19 is challenging, particularly in countries with limited healthcare system capacity and incomplete vital registration [29–31]. Consequently, some researchers prefer all-cause excess mortality, defined as the difference between observed and expected deaths, as a more reliable measure [6, 32]. In this study, I use both all-cause excess and COVID-19 mortality data.

All-cause mortality data were obtained from Peru’s National Information System of Deaths (SINADEF) [33]. An assessment concluded that SINADEF represents a substantial improvement over earlier death registries in Peru; although some incompleteness remains, no systematic difference in completeness by sex was observed [34]. In addition, COVID-19 case fatality data were analyzed [35]. These figures are subject to underreporting and have been subsequently adjusted by the authorities [30]. However, this subgroup was more likely affected by the CQ/HCQ treatment policy.

### Medicament data

I collected data on CQ/HCQ distributions from the database of the National Center for the Supply of Strategic Health Resources (CENARES) [36]. Since official treatment protocols treated CQ and HCQ as equivalents, I do not distinguish between them and summed their distributions after the conversion of CQ to HCQ-equivalents via protocol dosage ratios (see Table I, Appendix for details).

### Prevalence of G6PD deficiency

I used a map of Latin America provided by Monteiro et al. [28] showing model-based estimates of the prevalence of glucose-6-phosphate dehydrogenase deficiency (G6PDd). I overlaid the Peruvian state boundaries onto it (Figure V, Appendix) to assign prevalence values to each state (Table IV, Appendix).

### COVID-19 test results and vaccination data

I obtained COVID-19 test results from Peruvian authorities, which included both molecular (i.e., PCR) [37] and nonmolecular tests (i.e., rapid antibody tests, antigen tests, and chemiluminescence assays) [38]. Like official treatment protocols, I treated all types of tests equivalently. Additionally, I obtained COVID-19 vaccination data from Peruvian authorities [39] treating all vaccine types equivalently.

### Other Data

Data on the population were obtained from official statistics [40]. For reasons of feasibility, all rate calculations are based on the 2021 mid-year population estimates provided by the Ministry of Health of Peru [40], which readily include the necessary breakdowns by sex, age group, and region. Moreover, I consulted official documents containing CQ/HCQ treatment protocols for COVID-19 patients [9, 10, 41, 42].

## Methods

### Data preparation

Deaths categorized as violent (accidents, homicides, or suicides) and rare observations with missing information on sex, age, or location were excluded. All datasets were aggregated to the state level and to a weekly timeframe. The state of Callao is completely embedded within Lima and was therefore assigned to Lima instead of treating it separately. I stratified the data by the age groups 0–14, 15–44, 45–64, 65– 79, and 80+ and created the aggregated group 45+, which is the main focus of this analysis, as this group is considered to be most affected by COVID-19 and related treatments.

#### Coding of COVID-19 treatment protocols (recommendation_HCQ)

Official COVID-19 treatment protocols varied in their stance on CQ/HCQ [9, 10, 41, 42]. I coded these recommendations in an ordinal variable called *recommendation_HCQ* from 0 (no recommendation), to 1 (recommended only for hospitalized, i.e., medium and severe COVID-19 cases), 2 (additionally recommended for confirmed outpatient, i.e., mild cases), and 3 (additionally recommended for suspected outpatient, i.e., mild cases). Protocol changes were aligned with the weekly dataset by assigning each protocol to all calendar weeks between its issuance and the release of the subsequent update.

#### Definition of the CQ/HCQ exposure period (period_HCQ)

In line with the hypotheses, I constructed the binary variable *period_HCQ^2^* to indicate weeks when outpatient treatment with CQ/HCQ likely occurred and acute hemolytic anemia may have been mismanaged due to a strained healthcare system.

The variable *period_HCQ* was set to 1 if (i) the official COVID-19 treatment protocol recommended CQ/HCQ for outpatients, i.e., mild cases (*recommendation_HCQ* = 2 or 3), and (ii) the proxy for healthcare system strain and pandemic waves, i.e., the z-score of all-cause mortality (all ages) exceeded the threshold of 3 for at least two consecutive weeks. To account for potential delays between treatment and death, I extended the period by two additional weeks after the z-score went below the threshold.

In at least 11 states, however, CQ/HCQ was soon displaced by ivermectin (IVM) for outpatient treatment. In Piura, for example, the state health minister stated in early June 2020 (cw23) that HCQ for mild cases has been replaced by IVM owing to its perceived efficacy and fewer side effects [43]. Moreover, ten states (Ayacucho, Cajamarca, Cusco, Huancavelica, Huánuco, Junín, Moquegua, Pasco, Puno, Tacna) implemented mass IVM distributions through the Ministry of Defense’s Mega-Operación Tayta (MOT). During its first phase, MOT conducted household visits to identify vulnerable individuals; one week later, it began distributing IVM broadly, regardless of test status or symptoms, alongside other medications such as acetaminophen and azithromycin [44]. HCQ was not distributed as part of the MOT. Similarly, the regional government of Tumbes carried out a locally organized campaign with mass IVM distributions (Vanessa 2020a, 2020b). In these 11 states, I therefore shortened *period_HCQ* by ending it two weeks after the start of the mass IVM distribution to reflect a reasonable transition period.

Since *period_HCQ* is based on assumptions and reasoning, I reconstructed it under alternative definitions for robustness checks, using different z-score thresholds (2.5, 3, 3.5, 4) both with and without adjustments for IVM campaigns.

#### State-level CQ/HCQ availability (HCQ_per_pop)

Reliable state-level time series data on CQ/HCQ consumption are unavailable. Instead, I used data on the distribution of CQ/HCQ tablets to regional health authorities (i.e., stock data). For each state, I summed all distributions that occurred before or during *period_HCQ* and created a time-invariant variable, *HCQ_per_pop^3^*. To make the states comparable, I standardized the different types of drugs according to treatment protocols and divided the number of these tablets by the state population, expressed per 100,000 people (for details, see Table I, Appendix).

In most states, the quantities of distributed CQ/HCQ doses were limited relative to the total number of COVID-19 cases. On average, only 1.16 doses per positive test in the 45+ age group were delivered, decreasing to 0.4 doses when positive tests across all ages were considered (Table II, Appendix). Symptomatic but untested cases were also treated [42], indicating relative scarcity in most states and supporting the correspondence between stock data and actual usage. Moreover, self-medication with CQ/HCQ seems to have been uncommon in Peru [45–47], and web searches indicate limited public interest [48]. Therefore, *HCQ_per_pop* is unlikely to be strongly confounded by privately obtained doses. Although this measure does not capture actual intake or distinguish between inpatient and outpatient administration, *HCQ_per_pop* is expected to reasonably reflect interstate differences in CQ/HCQ availability.

### Statistical analysis

#### Estimating all-cause excess mortality rates

All-cause excess mortality is defined as the difference between the expected mortality and the observed mortality. I estimated weekly absolute numbers of expected deaths (baselines) for 2020–22 for each state, sex, and age group on the basis of historical data from 2017–19 via a time series regression model by Karlinsky and Kobak [12]. Absolute numbers of all-cause excess deaths were then calculated by subtracting the baseline from observed deaths, and crude mortality rates for each group were derived by dividing excess deaths by the corresponding population and scaling per 100,000 individuals. For feasibility and consistency, all rate calculations for 2020-22 used the 2021 mid-year population estimates provided by the Ministry of Health of Peru [40], which already include necessary breakdowns by sex, age group, and locality. This approach may not yield the most precise estimates of all-cause excess mortality rates. However, more sophisticated methods would likely provide little additional insight, as only three years of training data (2017-19) are available.

#### Age-standardized mortality rates (ASMR)

The confounding effect of different age structures makes comparisons between sexes for aggregated age groups misleading. Therefore, for the 45+ age group, I calculated the age-standardized mortality rates (ASMR) for males and females in each state via the direct method with Peru’s age structure as the standard population. The sex difference was then calculated as the difference in the ASMR between males and females. This was done separately for both all-cause excess and COVID-19 mortality rates.

To quantify the statistical uncertainty of the estimated all-cause excess ASMRs, I calculated 95% confidence intervals (CIs) using the predictive standard errors from the baseline mortality regressions. For each sex, the derived variances were summed across age groups using the squared standard-population weights. The CIs were constructed as the point estimate ± 1.96 times the square root of this variance.

#### TWFE model specification

The sex difference in all-cause excess, as well as in COVID-19 ASMR, for the 45+ age group was used as the dependent variable in two-way fixed-effects (TWFE) models, specified as follows:

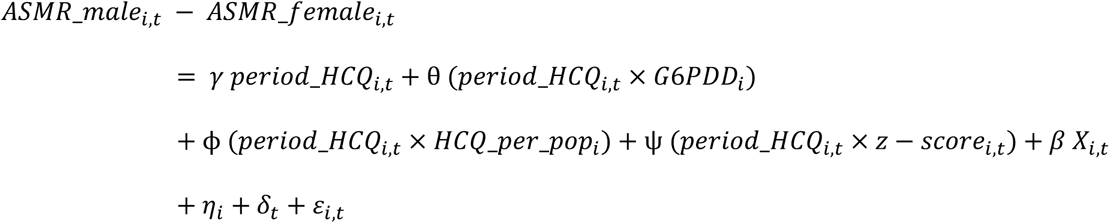

where *period_HCQ_i,t_* indicates the weeks with likely outpatient treatment with CQ/HCQ during a strained healthcare system in state *i* at time *t*, (*period_HCQ_i,t_* × *G6PDD_i_*), (*period_HCQ_i,t_* × *HCQ_per_pop_i_*), and (*period_HCQ_i,t_* × *z − score_i,t_*) are interaction terms capturing the conditional effects of G6PDd prevalence, QC/HCQ availability, and healthcare system strain—as measured by the all-cause mortality z-score for both sexes and all age groups—during period_HCQ, reflecting the hypotheses. *X_i,t_* is a vector of control variables, *η_i_* and *δ_t_* are state- and week-specific fixed effects, respectively, and *ε_i,t_* is the error term^4^. I used Driscoll-Kraay standard errors to account for cross-sectional dependence and serial correlation [49] and used wild cluster bootstrap inference [50] as a robustness check, given the small number of states.

The TWFE setup addresses several sources of confounding. First, time-invariant differences across states not included in the interaction terms are absorbed by the state fixed effects. Second, week fixed effects account for shocks common to all states at a given week. Third, using the sex difference in ASMR as the dependent variable implicitly removes factors that influence men and women in a similar way.

Control variables include the z-score, age-standardized sex differences in COVID-19 test positive rates, and cumulative sex differences in first-dose vaccination rates for individuals aged 45+ years. Analogous to the age standardization of mortality rates, test positivity rates were standardized via the national age distribution of tests to account for differences in testing by age. The cumulative vaccination rate sex differences were calculated as the sum over time of the male first-dose rates minus the female rates.

The z-score control variable estimates the ‘baseline’ relationship between overall excess mortality and sex differences in ASMR independent of *period_HCQ.* It captures the general male-biased mortality pattern observed during periods of increased all-cause mortality, whereas the z-score interaction term captures any additional effect during *period_HCQ*, reflecting the hypothesis that greater healthcare system strain could have further amplified sex differences in mortality by increasing the likelihood of adverse effects remaining unrecognized.

Because testing capacity in Peru was limited, particularly during the first wave, reported COVID-19 test results are an imperfect proxy for true infections. Consequently, the observed sex differences in test positivity may not fully capture actual infection patterns. Nonetheless, several considerations suggest that the resulting bias is limited. Nationwide lockdowns and contact restrictions were not gender-specific and are absorbed by week fixed effects, leaving only state-level reopening policies as a possible source of differential exposure. Moreover, household transmission was high—a study in Lima reported a secondary attack rate of 53% [51]—which likely reduced sustained disparities between sexes. Finally, Peruvian seroprevalence studies consistently show no significant sex differences in infection levels [52–54]. Overall, while short-term sex-specific fluctuations in infections may be partially captured by the control variable, persistent sex-specific differences in infection risk were small and unlikely to explain the observed sex difference in all-cause excess mortality.

## Results

### Sex differences during pandemic waves

Except for Apurímac, all Peruvian states recorded higher male than female ASMR in the 45+ age group during pandemic waves (i.e., periods ≥ 2 consecutive weeks with z-scores ≥ 3), leading to predominantly positive average sex differences in all-cause excess ASMR, as well as COVID-19 ASMR (Table 1). In most states, these differences were greater during the first pandemic wave (when CQ/HCQ was recommended) than in subsequent waves, with several cases showing confidence intervals that (nearly) exclude zero or p-values ≤0.05. Moreover, the size of the observed differences varied considerably, ranging from negligible or negative (Apurímac, Pasco) to very large (e.g., Madre de Dios, Moquegua, Ucayali).

**Table 1:** Average sex differences in all-cause excess and COVID-19 ASMR during pandemic waves.

| State | Weeks Wave 1 | Weeks Waves 2+ | Avg. all-cause excess sex diff. Wave 1 [95% CI] | Avg. all-cause excess sex diff. Waves 2+ [95% CI] | Diff. all-cause excess (W1–W2+) [95% CI] | COVID-19 Avg. Sex diff. Wave 1 | COVID-19 Avg. Sex diff. Waves 2+ | Diff. COVID-19 (W1–W2+) (p-value) |
| --- | --- | --- | --- | --- | --- | --- | --- | --- |
| Amazonas | 8 | 10 | 41.9 [18.3, 65.4] | 17.3 [-6.3, 40.8] | 24.6 [-8.7, 57.9] | 34.4 | 19.2 | 15.2 (0.040) |
| Ancash | 17 | 21 | 28.1 [12.6, 43.6] | 19.0 [3.5, 34.5] | 9.1 [-12.8, 31.1] | 24.2 | 18.4 | 5.8 (0.037) |
| Apurimac | 4 | 31 | -6.5 [-31.7, 18.8] | 2.9 [-22.3, 28.2] | -9.4 [-45.1, 26.4] | -1.1 | 5.4 | -6.6 (0.367) |
| Arequipa | 14 | 22 | 49.8 [36.0, 63.7] | 16.7 [2.8, 30.5] | 33.2 [13.6, 52.7] | 3.3 | 14.6 | -11.3 (0.078) |
| Ayacucho | 2 | 8 | 54.0 [28.4, 79.6] | 3.8 [-21.8, 29.4] | 50.2 [14.0, 86.4] | 27.2 | 8.9 | 18.3 (0.043) |
| Cajamarca | 10 | 20 | 24.8 [12.1, 37.5] | 11.9 [-0.9, 24.6] | 12.9 [-5.1, 30.9] | 19.8 | 12.3 | 7.5 (0.058) |
| Cusco | 6 | 18 | 33.4 [15.6, 51.2] | 10.8 [-6.9, 28.6] | 22.6 [-2.5, 47.7] | 7.4 | 14.7 | -7.4 (0.053) |
| Huancavelica | 6 | 18 | 30.1 [-8.5, 68.8] | 6.1 [-32.5, 44.8] | 24.0 [-30.7, 78.6] | 19.8 | 7.8 | 12.0 (0.030) |
| Huánuco | 12 | 35 | 12.8 [-2.9, 28.6] | 8.0 [-7.8, 23.7] | 4.8 [-17.4, 27.1] | 11.3 | 11.4 | -0.1 (0.966) |
| Ica | 23 | 30 | 36.6 [18.9, 54.4] | 21.3 [3.6, 39.1] | 15.3 [-9.8, 40.4] | 23.5 | 19.4 | 4.1 (0.337) |
| Junín | 11 | 23 | 32.4 [16.4, 48.4] | 25.7 [9.6, 41.7] | 6.7 [-15.9, 29.4] | 17.6 | 24.6 | -7.0 (0.152) |
| La Libertad | 18 | 25 | 30.2 [18.4, 42.1] | 9.9 [-1.9, 21.7] | 20.3 [3.6, 37.0] | 20.7 | 11.3 | 9.5 (0.010) |
| Lambayeque | 18 | 33 | 32.0 [11.6, 52.3] | 9.6 [-10.8, 29.9] | 22.4 [-6.4, 51.2] | 23.2 | 12.3 | 10.9 (0.004) |
| Lima + Callao | 27 | 31 | 28.7 [22.3, 35.0] | 12.9 [6.5, 19.2] | 15.8 [6.8, 24.8] | 20.7 | 16.5 | 4.2 (0.201) |
| Loreto | 14 | 15 | 58.8 [43.2, 74.4] | 18.2 [2.5, 33.8] | 40.6 [18.5, 62.7] | 63.0 | 15.0 | 48.0 (0.001) |
| Madre de Dios | 10 | 7 | 196.4 [131.4, 261.4] | 100.3 [35.3, 165.3] | 96.1 [4.2, 188.0] | 194.8 | 101.0 | 93.7 (0.012) |
| Moquegua | 10 | 12 | 119.3 [88.0, 150.5] | 19.4 [-11.9, 50.6] | 99.9 [55.7, 144.1] | 122.4 | 29.8 | 92.6 (0.000) |
| Pasco | 9 | 22 | 15.1 [-12.3, 42.6] | 19.9 [-7.6, 47.3] | -4.7 [-43.5, 34.1] | 11.0 | 11.9 | -0.9 (0.889) |
| Piura | 12 | 10 | 52.4 [34.6, 70.2] | 13.4 [-4.4, 31.3] | 39.0 [13.8, 64.2] | 45.8 | 20.6 | 25.2 (0.002) |
| Puno | 8 | 28 | 53.3 [39.9, 66.7] | 18.8 [5.4, 32.2] | 34.4 [15.5, 53.4] | 21.0 | 15.1 | 5.8 (0.084) |
| San Martin | 12 | 16 | 41.8 [21.3, 62.2] | 18.1 [-2.3, 38.6] | 23.6 [-5.3, 52.5] | 40.4 | 18.3 | 22.1 (0.002) |
| Tacna | 10 | 25 | 71.0 [42.7, 99.4] | 35.1 [6.8, 63.5] | 35.9 [-4.2, 76.0] | 57.2 | 27.9 | 29.2 (0.012) |
| Tumbes | 11 | 17 | 58.1 [22.2, 94.1] | 36.8 [0.8, 72.7] | 21.4 [-29.4, 72.2] | 54.7 | 42.0 | 12.7 (0.238) |
| Ucayali | 6 | 9 | 110.0 [85.2, 134.7] | 39.1 [14.3, 63.8] | 70.9 [35.9, 105.9] | 95.3 | 50.7 | 44.6 (0.017) |
| Median | 10.5 | 20.5 | 41.8 | 17.8 | 23.6 | 23.4 | 15.8 | 10.2 |
| Mean | 11.6 | 20.3 | 50.2 | 21.3 | 28.9 | 39.2 | 21.8 | 17.5 |
| Std. Dev. | 5.6 | 8.1 | 39.9 | 18.5 | 26.4 | 42.1 | 19.0 | 26.5 |

Note: Average ASMR sex differences (males minus females) for individuals aged 45+ in deaths per 100,000. Waves are defined as periods of $\geq 2$ consecutive weeks with a z-score $\geq 3$ . The
95% confidence intervals (95% CIs) are based on the predictive variance of the baseline models. P-values are from two-sided permutation tests.

Figure 1 shows the male and female all-cause excess ASMR for those aged 45+ over time, with markedly higher values for males, especially during the 2020 wave, and a close correspondence with the all-age mortality z-score. There is no period where female all-cause excess mortality significantly exceeds male mortality (i.e., a negative sex difference) and regional heterogeneity is evident. Periods where *period_HCQ* =1 (i.e., weeks of substantial healthcare system strain (z-score ≥3) combined with the CQ/HCQ recommendation for COVID-19 outpatients) are also indicated. Comparable figures for all-cause sex differences and for the COVID-19 ASMR are provided in the Appendix (Figure I, Figure II, Appendix).

**Figure 1:**
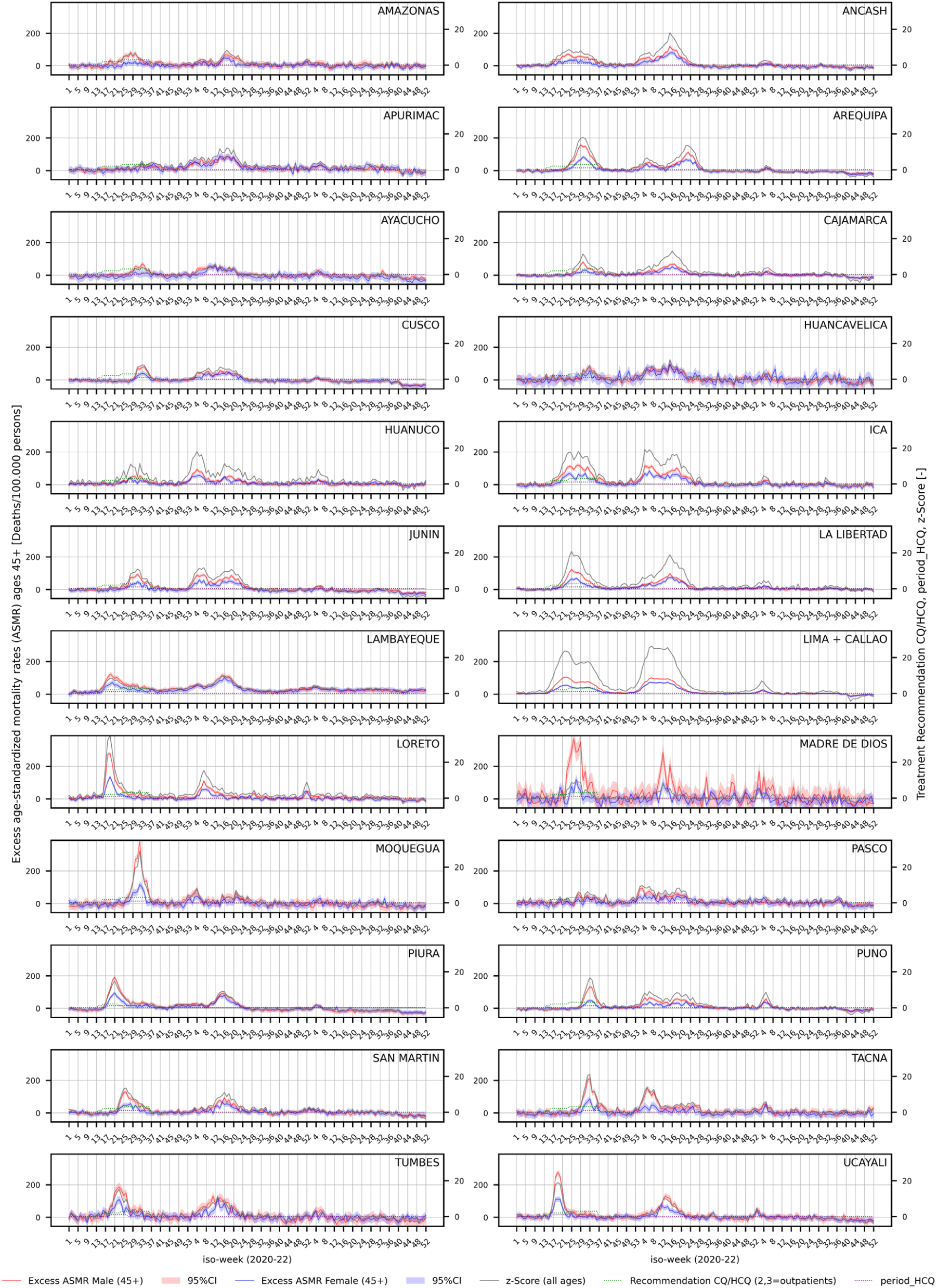
Male and female all-cause excess ASMR for individuals aged 45+ over time by state. Note: 95% CI stands for 95% confidence 261 interval.

### TWFE model results

I present two models for each of the two outcomes (all-cause excess, COVID-19) to separate the main and interaction effects. In models 2 and 4 (Table 2), the main effect of *period_HCQ* should be interpreted with caution, since it is not only identified at unrealistic values of the interactions (all zero), but is also inflated by multicollinearity with the interaction terms. Models 1 and 3 (Table 2) show a positive and significant coefficient for *period_HCQ* for both outcome variants (22.049, p ≤ 0.001; 15.689, p ≤ 0.001), indicating that the difference between male and female ASMR increased during the period when CQ/HCQ was officially recommended for COVID-19 outpatients and the healthcare system was strained (z-score ≥ 3). However, this allows no conclusions regarding the hypothesized mechanism because it does not disentangle the contributions of G6PDd prevalence, regional HCQ availability, or intensity of healthcare system strain.

**Table 2:**
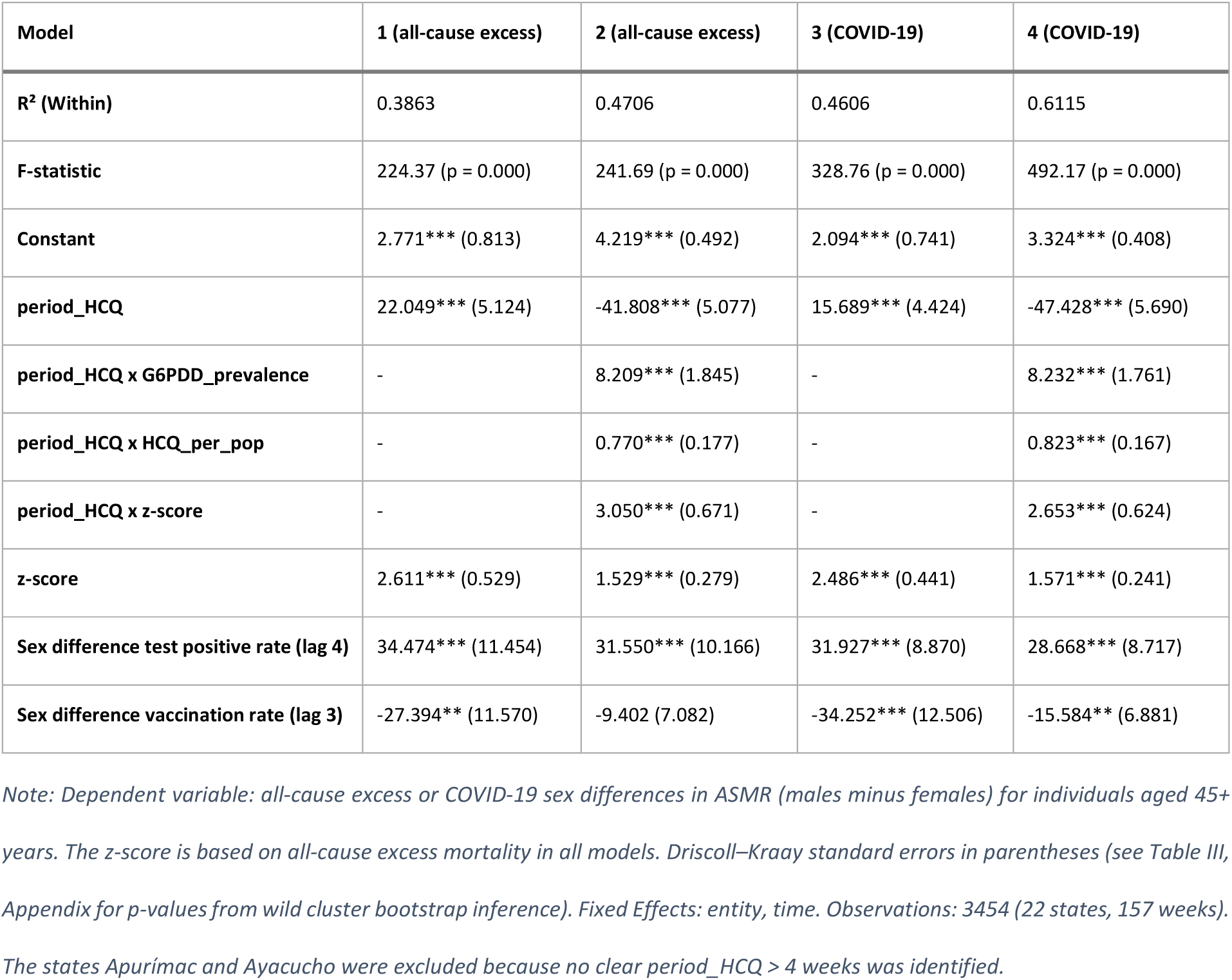
Results of the TWFE models.

| Model | 1 (all-cause excess) | 2 (all-cause excess) | 3 (COVID-19) | 4 (COVID-19) |
| --- | --- | --- | --- | --- |
| **R<sup>2</sup> (Within)** | 0.3863 | 0.4706 | 0.4606 | 0.6115 |
| **F-statistic** | 224.37 (p = 0.000) | 241.69 (p = 0.000) | 328.76 (p = 0.000) | 492.17 (p = 0.000) |
| **Constant** | 2.771*** (0.813) | 4.219*** (0.492) | 2.094*** (0.741) | 3.324*** (0.408) |
| **period_HCQ** | 22.049*** (5.124) | -41.808*** (5.077) | 15.689*** (4.424) | -47.428*** (5.690) |
| **period_HCQ x G6PDD_prevalence** | - | 8.209*** (1.845) | - | 8.232*** (1.761) |
| **period_HCQ x HCQ_per_pop** | - | 0.770*** (0.177) | - | 0.823*** (0.167) |
| **period_HCQ x z-score** | - | 3.050*** (0.671) | - | 2.653*** (0.624) |
| **z-score** | 2.611*** (0.529) | 1.529*** (0.279) | 2.486*** (0.441) | 1.571*** (0.241) |
| **Sex difference test positive rate (lag 4)** | 34.474*** (11.454) | 31.550*** (10.166) | 31.927*** (8.870) | 28.668*** (8.717) |
| **Sex difference vaccination rate (lag 3)** | -27.394** (11.570) | -9.402 (7.082) | -34.252*** (12.506) | -15.584** (6.881) |

Note: Dependent variable: all-cause excess or COVID-19 sex differences in ASMR (males minus females) for individuals aged 45+ years. The z-score is based on all-cause excess mortality in all models. Driscoll–Kraay standard errors in parentheses (see Table III, Appendix for p-values from wild cluster bootstrap inference). Fixed Effects: entity, time. Observations: 3454 (22 states, 157 weeks). The states Apurímac and Ayacucho were excluded because no clear period\_HCQ > 4 weeks was identified.

Models 2 and 4 introduce interaction terms with G6PDd prevalence, regional CQ/HCQ availability, and the all-cause *z-score*. This specification allows the effect of *period_HCQ* to vary by regional risk factors and healthcare system strain (proxied by the *z-score*), effectively isolating the hypothesized mechanism. All three interactions are positive and highly significant for both outcome variants: *period_HCQ × G6PDd prevalence* (8.209, p ≤0.001; 8.232, p ≤0.001), *period_HCQ × HCQ_per_pop* (0.770, p ≤0.001; 0.823, p ≤0.001), and *period_HCQ × z-score* (3.050, p ≤0.001; 2.653, p ≤0.001). For example, a one-category increase in G6PDd prevalence – corresponding to a two-percentage-point increase (Table IV, Appendix) – alters the effect of *period_HCQ* by an additional sex difference of 8.209 all-cause excess deaths or 8.232 COVID-19 deaths per 100,000. Marginal effects plots for G6PDd prevalence are provided in the appendix (Figures III and IV). The coefficients’ magnitudes of the interaction terms should be interpreted with caution, as they vary slightly over the tested definitions of *period_HCQ.* However, significance and signs are robust and consistent with the tested mechanism.

Taken together, the results of the interaction terms suggest that in regions with higher G6PDd prevalence, greater CQ/HCQ availability, and stronger healthcare system strain, sex differences in all-cause excess and COVID-19 ASMR increased disproportionately during the CQ/HCQ treatment policy for outpatients, supporting the hypothesized mechanism of drug-induced hemolytic anemia under stressed healthcare system conditions.

The control variables also behaved as expected: the *z-score* captures the general male-biased mortality pattern observed during periods of increased all-cause mortality and is positive and significant. The coefficient of the 4-week lagged sex difference in test positive rates formally reflects a 100% difference (i.e., only men tested positive and no women) and is positive and significant in all models. It should be interpreted as capturing short-term shifts in infection burdens between sexes, as discussed in the Methods section. Although this variable is an imperfect proxy for true infection rates, its significance reflects that observed mortality patterns overlapped with periods and regions in which one sex was more likely to be registered as infected.

The 3-week lagged sex difference in first-dose vaccination rates has a negative coefficient, suggesting that weeks with relatively higher rates of female vaccination are associated with smaller sex differences in mortality. As with the test-positivity variable, the coefficient formally reflects a 100% difference and should be interpreted as capturing relative shifts in vaccination coverage between the sexes during the nationwide vaccination campaign starting in April 2021. The effect is somewhat unstable across specifications and was observed only in 2021, so it should be interpreted cautiously. While it provides some insight into sex-differential dynamics during the vaccination campaign, this variable does not contribute to the identification of *period_HCQ* and serves as a supplementary result.

To further assess which sex drives the effect of increased sex differences in ASMR during *period_HCQ*, separate models were estimated with male and female ASMR as outcomes. While these specifications are subject to omitted variable bias and endogeneity, they serve as a diagnostic check and show a strong effect for males but only negligible and mostly insignificant effects for females, suggesting that the observed sex differences stem from higher male mortality rather than unusually low female mortality.

## Discussion

The findings from Peru align with those of prior studies showing among men higher COVID-19 mortality [4, 5] and all-cause excess mortality [3]. Significant periods with higher female than male all-cause excess mortality, as observed in Thailand [6], were not present in Peru. This difference can be explained partly by the exclusion of traffic accidents in this analysis, which were probably responsible for the observed pattern in Thailand [6]. The correlation between general excess mortality (z-score) and sex differences, as found in Europe [3], was also confirmed by this analysis.

Moreover, the results are consistent with those of previous studies linking CQ/HCQ to increased all-cause mortality [13], especially in the Peruvian context [14].

Except for some case reports [22–26], the risk of CQ/HCQ-induced acute hemolytic anemia in G6PD-deficient patients is generally considered low according to a systematic review of the available evidence [21]. This study, however, suggests that this risk may have been underestimated in the specific context of Peru during the COVID-19 crisis, which was characterized by panic over an unknown virus, nontransparent treatment protocols [55], misuse of medications [56, 57], and an overburdened healthcare system. Under such circumstances, uncontrolled outpatient use of CQ and HCQ may have exposed G6PD-deficient individuals to increased hemolytic risk, potentially contributing to the observed male–female mortality gap. This hypothesis warrants further investigation using individual-level data linking medication use, hematologic indicators, and G6PD status.

The Peruvian prison system may be a suitable sample for future research. From an ecological perspective, a temporal association between the CQ/HCQ distribution and male mortality was also observed there. Until October 2020, 445 Peruvian prison inmates have died from COVID-19, 251 (56%) of whom died between April and May [58], which correlates with the delivery of 10,800 tablets of 400 mg HCQ in mid-April [36].

There are several limitations to this study that are worth mentioning. First, the model is ecological in nature and cannot establish individual-level causality. Second, the state-level measure of G6PDd prevalence [28] is modeled and relatively coarse, which may blunt interaction effects. Third, the measure of CQ/HCQ availability is based on stock data per population and does not capture actual intake, compliance, informal distribution, black-market flows, or tablets sold via the private health sector. Fourth, the z-score of the all-cause ASMR serves only as an indirect indicator of healthcare system strain and does not reflect ICU capacity, staffing shortages, etc. for which no data is available. Finally, the all-cause excess ASMR were predicted by the model by Karlinsky and Kobak [12] and were used directly in the TWFE analysis without formally accounting for estimation uncertainty, which may have resulted in underestimated standard errors.

## Conclusion

This study indicates that Peru’s nationwide CQ/HCQ treatment policy during the early COVID-19 pandemic may have contributed to sex-specific differences in mortality among adults aged 45+, particularly in regions with higher glucose-6-phosphate dehydrogenase deficiency (G6PDd) prevalence and during periods of elevated healthcare system strain. Across states, both male all-cause excess and COVID-19 mortality consistently exceeded female mortality, with the largest male‒female differences coinciding with periods of plausible widespread CQ/HCQ use among COVID-19 outpatients. Interaction analyses indicate that regions with higher G6PDd prevalence, greater CQ/HCQ availability, and elevated all-age excess mortality (proxying healthcare system strain) experienced amplified male-female mortality gaps.

These findings are consistent with the hypothesis that CQ/HCQ-induced oxidative stress may have triggered hemolytic crises in (undiagnosed) G6PDd patients. In the context of COVID-19, where infection itself causes oxidative stress and potentially hemolysis [27] and presents with overlapping symptoms (e.g., dyspnea, fever), such adverse reactions may have been overlooked or misclassified as severe COVID-19 in overstretched healthcare systems.

While causality cannot be established owing to the ecological study design, these findings underscore the importance of incorporating genetic and sex-specific risk factors into pandemic treatment guidelines, particularly in resource-limited settings with high uncertainty. The oxidative stress induced by COVID-19 infection represents a distinct clinical context, in which drugs previously considered safe — such as CQ/HCQ for widespread malaria treatment — may pose unforeseen risks for G6PD-deficient individuals. Given the likelihood that hemolysis may have been unrecognized, ecological analyses remain crucial for detecting population-level associations and generating hypotheses for future research and policy-making.

## Data Availability

All data produced in the present study are available upon reasonable request to the authors

## Declarations

### Ethics approval and consent to participate

Not applicable

### Consent for publication

Not applicable

### Availability of data and materials

The datasets used and/or analysed during the current study are available from the corresponding author on reasonable request.

### Authors’ contributions

MK conceived the study, collected and analyzed the data, interpreted the results, and wrote the manuscript. The author read and approved the final manuscript.

## Acknowledgements

Not applicable

## Appendix

**Figure I:**
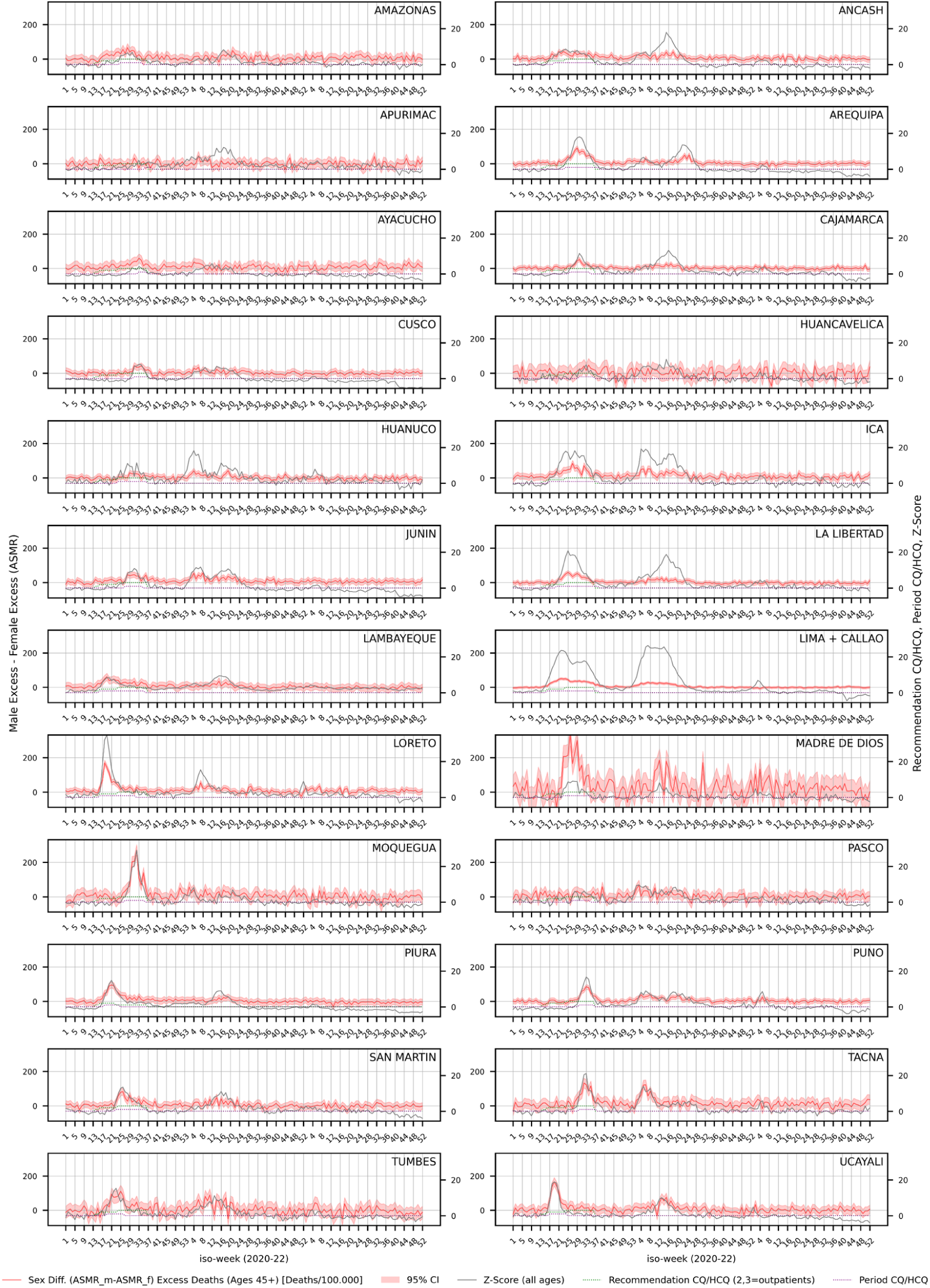
Sex differences in all-cause excess ASMR (males-females) for individuals aged 45+ over time by state *Note: The variance of the sex difference was obtained as the sum of male and female ASMR variances (see the* Statistical Analysis section), *and the 95% confidence interval (95% CI) was constructed as the point estimate ± 1.96 times the square root of this variance*, *assuming the independence of errors (Cov(m*, *f) = 0)*.

**Figure II:**
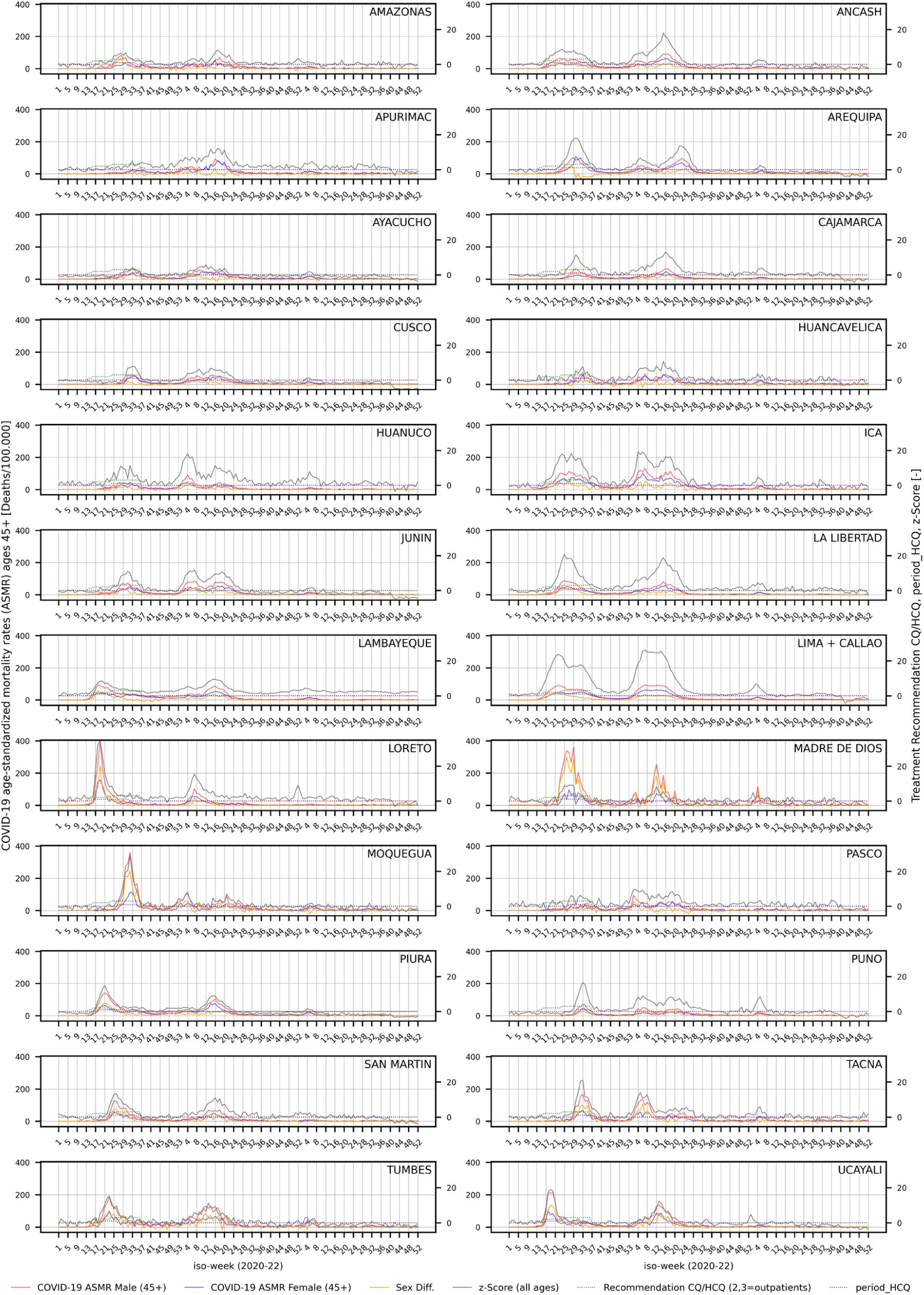
Sex differences in COVID-19 ASMR (males-females) for individuals aged 45+ over time by state

**Table I:**
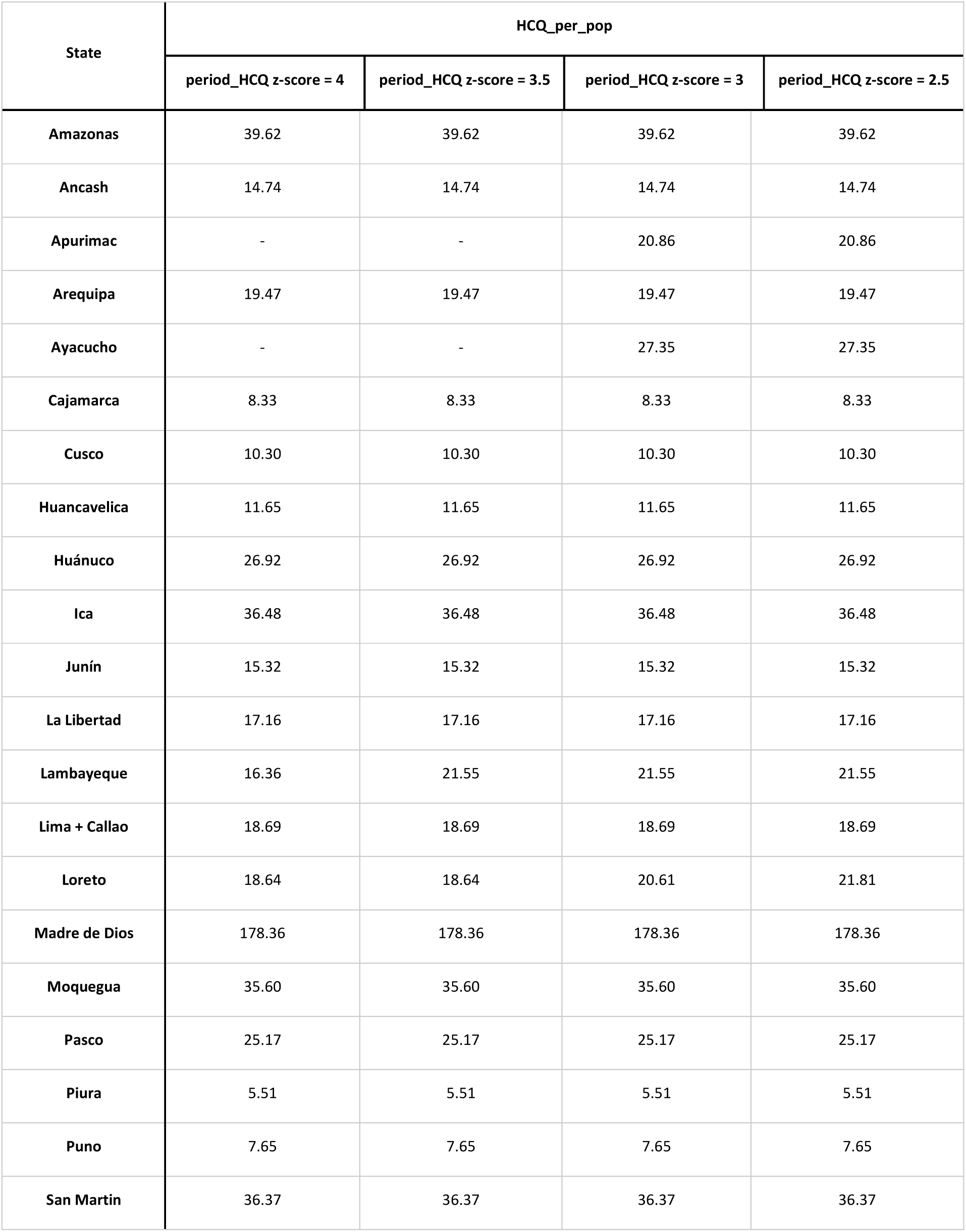
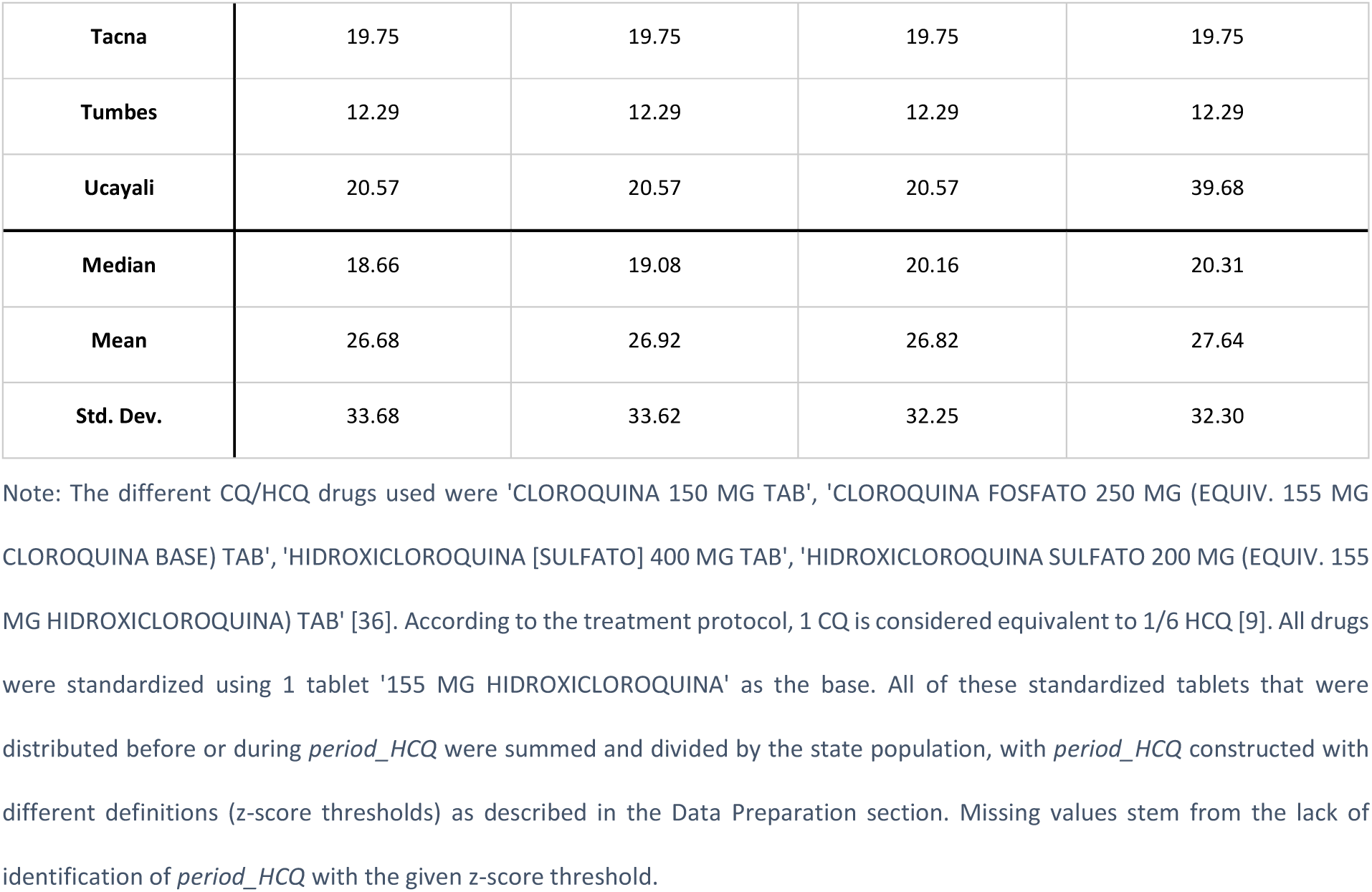
Sum of delivered HCQ tablets per state population.

**Table II:** Scarcity CQ/HCQ: Doses per positive test.

| State | CQ/HCQ-Doses per positive tests in age group 45+ |  |  |  | Total tests per population during Period_HCQ (z-score = 2.5) |
| --- | --- | --- | --- | --- | --- |
|  | period_HCQ z-score = 4 | period_HCQ z-score = 3.5 | period_HCQ z-score = 3 | period_HCQ z-score = 2.5 |  |
| **Amazonas** | 1.36 | 1.21 | 1.21 | 1.11 | 0.19 |
| **Ancash** | 0.46 | 0.46 | 0.42 | 0.41 | 0.21 |
| **Apurimac** | NaN | NaN | 4.48 | 2.12 | 0.13 |
| **Arequipa** | 0.23 | 0.22 | 0.22 | 0.22 | 0.50 |
| **Ayacucho** | NaN | NaN | 3.30 | 2.11 | 0.06 |
| **Cajamarca** | 0.59 | 0.59 | 0.57 | 0.57 | 0.10 |
| Cusco | 0.41 | 0.41 | 0.41 | 0.41 | 0.14 |
| Huancavelica | 1.36 | 1.09 | 1.09 | 1.09 | 0.09 |
| Huánuco | 1.23 | 1.14 | 1.14 | 1.14 | 0.14 |
| Ica | 0.54 | 0.54 | 0.54 | 0.54 | 0.39 |
| Junín | 1.12 | 1.03 | 1.03 | 1.03 | 0.10 |
| La Libertad | 0.51 | 0.51 | 0.48 | 0.48 | 0.19 |
| Lambayeque | 0.94 | 0.80 | 0.70 | 0.59 | 0.17 |
| Lima + Callao | 0.27 | 0.27 | 0.27 | 0.27 | 0.40 |
| Loreto | 1.38 | 1.38 | 1.09 | 0.94 | 0.08 |
| Madre de Dios | 3.66 | 3.35 | 3.35 | 3.05 | 0.26 |
| Moquegua | 0.40 | 0.40 | 0.39 | 0.39 | 0.48 |
| Pasco | 1.80 | 1.68 | 1.68 | 1.68 | 0.14 |
| Piura | 0.64 | 0.64 | 0.64 | 0.64 | 0.04 |
| Puno | 0.56 | 0.56 | 0.56 | 0.56 | 0.08 |
| San Martín | 1.36 | 1.36 | 1.29 | 1.29 | 0.11 |
| Tacna | 0.51 | 0.51 | 0.51 | 0.49 | 0.26 |
| Tumbes | 1.01 | 1.01 | 1.01 | 1.01 | 0.05 |
| Ucayali | 1.80 | 1.80 | 1.80 | 2.17 | 0.07 |
| Median | 0.79 | 0.72 | 0.85 | 0.79 | 0.14 |
| Mean | 1.00 | 0.94 | 1.16 | 1.00 | 0.18 |
| Std. Dev. | 0.73 | 0.67 | 1.04 | 0.70 | 0.13 |

Note: CQ/HCQ deliveries [36] that occurred before or during *period\_HCQ* were considered, and 1 CQ/HCQ dose corresponds to 18 tablets of HIDROXICLOROQUINA SULFATO 200 MG [41]. All positive COVID-19 tests in the 45+ age group that occurred during

**Table III:** Inference with Wild Cluster Bootstrap [50] for Models 1, 2, 3, and 4 (Table 2, main text)

| Model | p-value (Wild Cluster Bootstrap) |  |  |  |
| --- | --- | --- | --- | --- |
|  | 1 (all-cause excess) | 2 (all-cause excess) | 3 (COVID-19) | 4 (COVID-19) |
| period_HCQ | 0.000 | 0.001 | 0.002 | 0.000 |
| period_HCQ x G6PDD prevalence | - | 0.075 | - | 0.023 |
| period_HCQ x HCQ_per_pop | - | 0.046 | - | 0.003 |
| period_HCQ x z-score | - | 0.000 | - | 0.004 |
| z-score | 0.000 | 0.000 | 0.000 | 0.000 |
| Sex difference test positive rate (lag 4) | 0.028 | 0.029 | 0.003 | 0.003 |
| Sex difference vaccination rate (lag 3) | 0.002 | 0.051 | 0.000 | 0.007 |

**Figure III:**
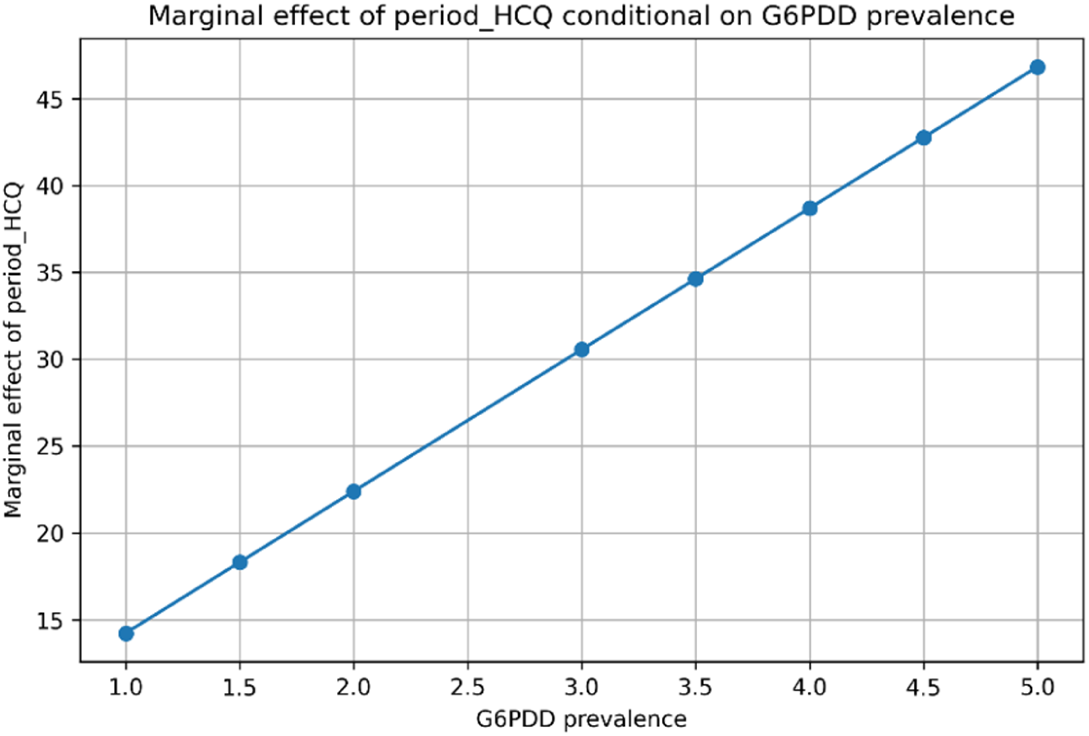
Marginal effect of period_HCQ conditional on G6PDd prevalence. Dependent variable: sex difference in all-cause excess ASMR (males-females) at age 45+ Note: The variable *HCQ_per_pop* (time-invariant) is held at its mean. *The variable z-score* is held at its mean during *period_HCQ*.

**Figure IV:**
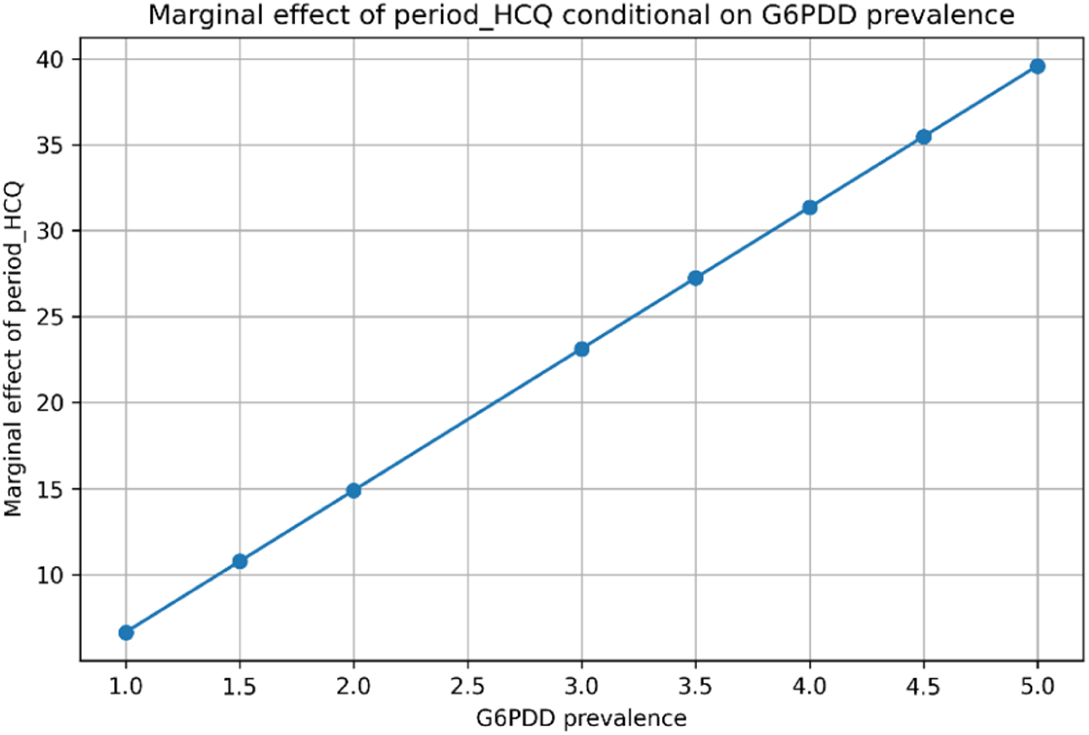
Marginal effect of period_HCQ conditional on G6PDd prevalence. Dependent variable: sex difference in Covid-19 ASMR (males-females) ages 45+ Note: The variable *HCQ_per_pop* (time-invariant) is held at its mean. *The variable z-score* is held at its mean during *period_HCQ*.

**Figure V:**
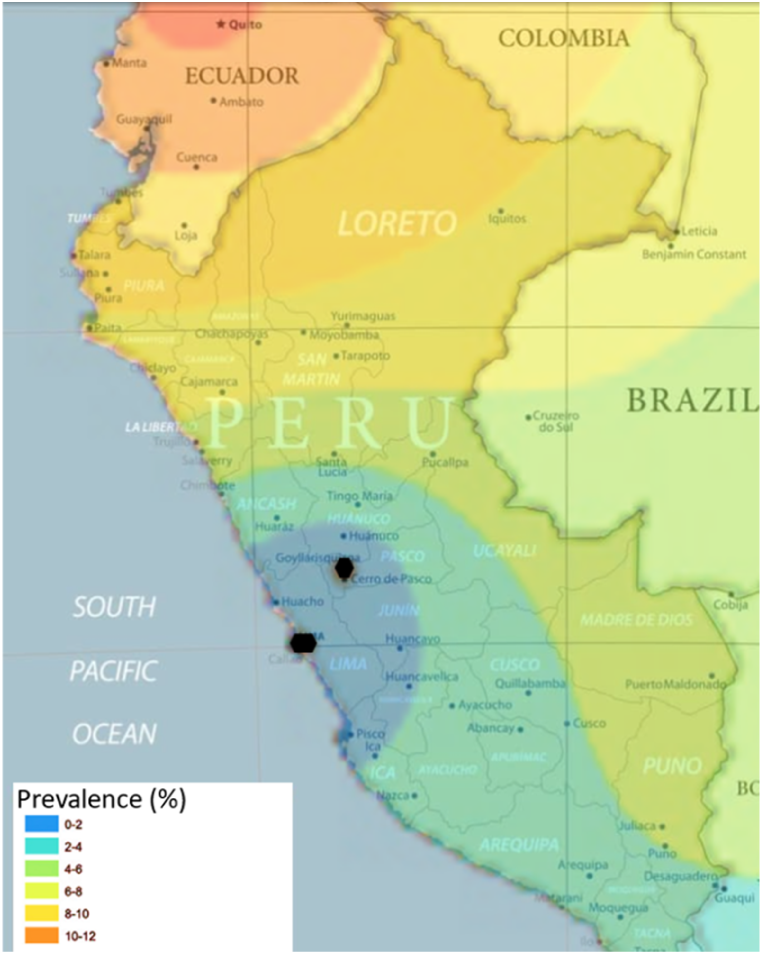
States of Peru and modeled G6PDd prevalence [28]

**Table IV:**
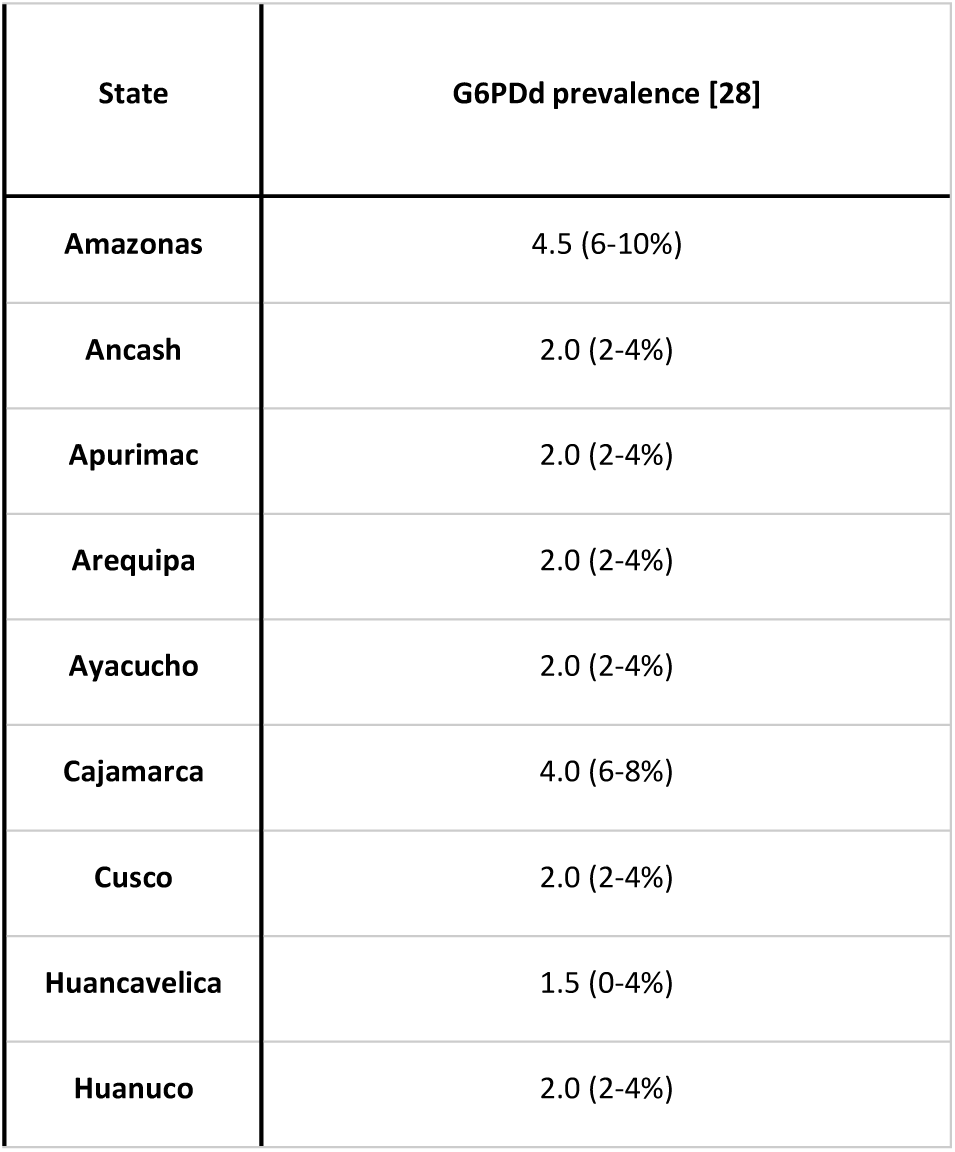
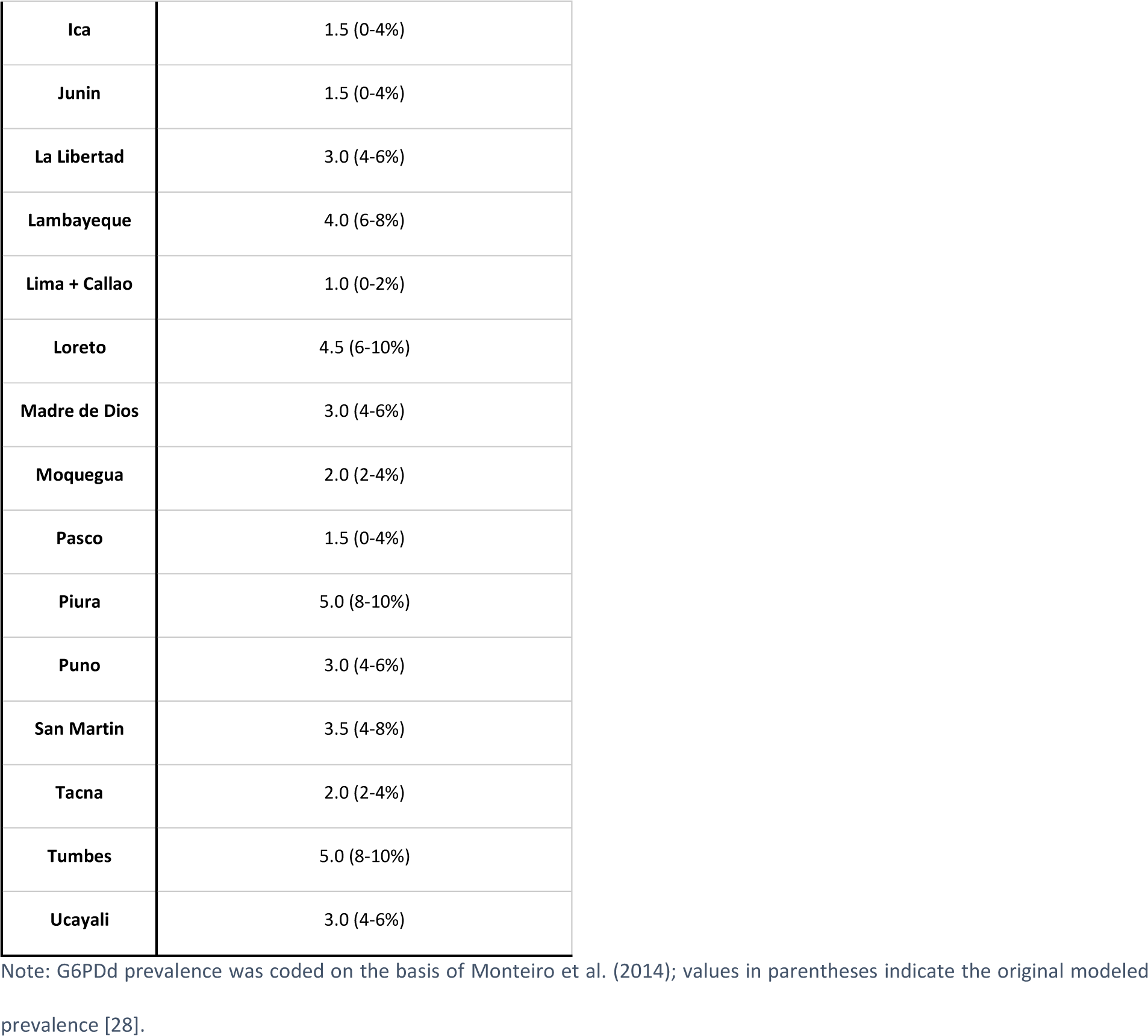
G6PDd prevalence by state, derived from [28].

| State | G6PDd prevalence [28] |
| --- | --- |
| Amazonas | 4.5 (6-10%) |
| Ancash | 2.0 (2-4%) |
| Apurimac | 2.0 (2-4%) |
| Arequipa | 2.0 (2-4%) |
| Ayacucho | 2.0 (2-4%) |
| Cajamarca | 4.0 (6-8%) |
| Cusco | 2.0 (2-4%) |
| Huancavelica | 1.5 (0-4%) |
| Huanuco | 2.0 (2-4%) |
| **Ica** | 1.5 (0-4%) |
| **Junin** | 1.5 (0-4%) |
| **La Libertad** | 3.0 (4-6%) |
| **Lambayeque** | 4.0 (6-8%) |
| **Lima + Callao** | 1.0 (0-2%) |
| **Loreto** | 4.5 (6-10%) |
| **Madre de Dios** | 3.0 (4-6%) |
| **Moquegua** | 2.0 (2-4%) |
| **Pasco** | 1.5 (0-4%) |
| **Piura** | 5.0 (8-10%) |
| **Puno** | 3.0 (4-6%) |
| **San Martin** | 3.5 (4-8%) |
| **Tacna** | 2.0 (2-4%) |
| **Tumbes** | 5.0 (8-10%) |
| **Ucayali** | 3.0 (4-6%) |

Note: G6PDd prevalence was coded on the basis of Monteiro et al. (2014); values in parentheses indicate the original modeled prevalence [28].

## Footnotes

2 For simplicity, I use here ‘HCQ’ to refer to both chloroquine and hydroxychloroquine.

3 For simplicity, I use ‘HCQ’ to refer to both chloroquine and hydroxychloroquine.

4 For simplicity, I use ‘HCQ’ in the variable names to refer to both chloroquine and hydroxychloroquine.

## References

1. Peretz C, Rotem N, Keinan-Boker L, Furshpan A, Green M, Bitan M, Steinberg DM. Excess mortality in Israel associated with COVID-19 in 2020-2021 by age group and with estimates based on daily mortality patterns in 2000-2019. Int J Epidemiol. 2022;51:727–36. doi:10.1093/ije/dyac047.

2. Henry NJ, Elagali A, Nguyen M, Chipeta MG, Moore CE. Variation in excess all-cause mortality by age, sex, and province during the first wave of the COVID-19 pandemic in Italy. Sci Rep. 2022;12:1077. doi:10.1038/s41598-022-04993-7.

3. Nielsen J, Nørgaard SK, Lanzieri G, Vestergaard LS, Moelbak K. Sex-differences in COVID-19 associated excess mortality is not exceptional for the COVID-19 pandemic. Sci Rep. 2021;11:20815. doi:10.1038/s41598-021-00213-w.

4. Bambra C, Albani V, Franklin P. COVID-19 and the gender health paradox. Scand J Public Health. 2021;49:17–26. doi:10.1177/1403494820975604.

5. Peckham H, Gruijter NM de, Raine C, Radziszewska A, Ciurtin C, Wedderburn LR, et al. Male sex identified by global COVID-19 meta-analysis as a risk factor for death and ITU admission. Nat Commun. 2020;11:6317. doi:10.1038/s41467-020-19741-6.

6. Pothisiri W, Prasitsiriphon O, Apakupakul J, Ploddi K. Gender differences in estimated excess mortality during the COVID-19 pandemic in Thailand. BMC Public Health. 2023;23:1900. doi:10.1186/s12889-023-16828-9.

7. Akter S. The Gender Gap in COVID-19 Mortality in the United States. Feminist Economics. 2021;27:30–47. doi:10.1080/13545701.2020.1829673.

8. World Health Organization. WHO discontinues hydroxychloroquine and lopinavir/ritonavir treatment arms for COVID-19. 2020. https://www.who.int/news/item/04-07-2020-who-discontinues-hydroxychloroquine-and-lopinavir-ritonavir-treatment-arms-for-covid-19. Accessed 4 Aug 2025.

9. Ministerio de Salud (MINSA). Resolución Ministerial N.° 139-2020-MINSA; 2020.

10. Ministerio de Salud (MINSA). Resolución Ministerial N.° 193-2020-MINSA; 2020.

11. Gestión. Minsa anuncia que la hidroxicloroquina se retirará de la guía de medicamentos para el COVID-19. 2020. https://gestion.pe/peru/coronavirus-peru-pilar-mazzetti-anuncia-que-la-hidroxicloroquina-se-retirara-de-la-guia-de-medicamentos-para-tratamiento-del-covid-19-nndc-noticia/. Accessed 4 Sep 2025.

12. Karlinsky A, Kobak D. Tracking excess mortality across countries during the COVID-19 pandemic with the World Mortality Dataset. Elife 2021. doi:10.7554/eLife.69336.

13. Axfors C, Schmitt AM, Janiaud P, Van’t Hooft J, Abd-Elsalam S, Abdo EF, et al. Mortality outcomes with hydroxychloroquine and chloroquine in COVID-19 from an international collaborative meta-analysis of randomized trials. Nat Commun. 2021;12:2349. doi:10.1038/s41467-021-22446-z.

14. Soto-Becerra P, Culquichicón C, Hurtado-Roca Y, Araujo-Castillo RV. Real-world effectiveness of hydroxychloroquine, azithromycin, and ivermectin among hospitalized COVID-19 patients: results of a target trial emulation using observational data from a nationwide healthcare system in Peru; 2020.

15. Mokhtari M, Mohraz M, Gouya MM, Namdari Tabar H, Tabrizi J-S, Tayeri K, et al. Clinical outcomes of patients with mild COVID-19 following treatment with hydroxychloroquine in an outpatient setting. Int Immunopharmacol. 2021;96:107636. doi:10.1016/j.intimp.2021.107636.

16. Lofgren SM, Nicol MR, Bangdiwala AS, Pastick KA, Okafor EC, Skipper CP, et al. Safety of Hydroxychloroquine Among Outpatient Clinical Trial Participants for COVID-19. Open Forum Infect Dis. 2020;7:ofaa500. doi:10.1093/ofid/ofaa500.

17. Lagier J-C, Million M, Gautret P, Colson P, Cortaredona S, Giraud-Gatineau A, et al. Outcomes of 3,737 COVID-19 patients treated with hydroxychloroquine/azithromycin and other regimens in Marseille, France: A retrospective analysis. Travel Med Infect Dis. 2020;36:101791. doi:10.1016/j.tmaid.2020.101791.

18. Million M, Lagier J-C, Gautret P, Colson P, Fournier P-E, Amrane S, et al. Early treatment of COVID-19 patients with hydroxychloroquine and azithromycin: A retrospective analysis of 1061 cases in Marseille, France. Travel Med Infect Dis. 2020;35:101738. doi:10.1016/j.tmaid.2020.101738.

19. Mohana A, Sulaiman T, Mahmoud N, Hassanein M, Alfaifi A, Alenazi E, et al. Hydroxychloroquine Safety Outcome within Approved Therapeutic Protocol for COVID-19 Outpatients in Saudi Arabia. Int J Infect Dis. 2021;102:110–4. doi:10.1016/j.ijid.2020.10.031.

20. Luzzatto L, Nannelli C, Notaro R. Glucose-6-Phosphate Dehydrogenase Deficiency. Hematol Oncol Clin North Am. 2016;30:373–93. doi:10.1016/j.hoc.2015.11.006.

21. Kane M. Hydroxychloroquine Therapy and G6PD Genotype. 2023. https://www.ncbi.nlm.nih.gov/books/NBK591356/. Accessed 4 Sep 2025.

22. Aguilar J, Averbukh Y. Hemolytic Anemia in a Glucose-6-Phosphate Dehydrogenase-Deficient Patient Receiving Hydroxychloroquine for COVID-19: A Case Report. Perm J 2020. doi:10.7812/TPP/20.158.

23. Sgherza N, Dalfino L, Palma A, Vitucci A, Campanale D, Grasso S, Musto P. “Hemolysis, or not Hemolysis, that is the Question”. Use of Hydroxychloroquine in a Patient with COVID-19 Infection and G6PD Deficiency. Mediterr J Hematol Infect Dis. 2020;12:e2020076. doi:10.4084/MJHID.2020.076.

24. Chaney S, Basirat A, McDermott R, Keenan N, Moloney E. COVID-19 and hydroxychloroquine side-effects: glucose 6-phosphate dehydrogenase deficiency (G6PD) and acute haemolytic anaemia. QJM. 2020;113:890–1. doi:10.1093/qjmed/hcaa267.

25. Mastroianni F, Colombie V, Claes G, Gilles A, Vandergheynst F, Place S. Hydroxychloroquine in a G6PD-Deficient Patient with COVID-19 Complicated by Haemolytic Anaemia: Culprit or Innocent Bystander? Eur J Case Rep Intern Med. 2020;7:1875. doi:10.12890/2020_001875.

26. Maillart E, Leemans S, van Noten H, Vandergraesen T, Mahadeb B, Salaouatchi MT, et al. A case report of serious haemolysis in a glucose-6-phosphate dehydrogenase-deficient COVID-19 patient receiving hydroxychloroquine. Infect Dis (Lond). 2020;52:659–61. doi:10.1080/23744235.2020.1774644.

27. Ayyad M, Abu Alya W, Barabrah AM, Darawish SM, AlHabil Y, MohammedAli M, et al. Autoimmune hemolytic anemia in COVID-19 patients: A systematic review of 105 cases on clinical characteristics and outcomes. Clin Immunol. 2025;277:110512. doi:10.1016/j.clim.2025.110512.

28. Monteiro WM, Val FFA, Siqueira AM, Franca GP, Sampaio VS, Melo GC, et al. G6PD deficiency in Latin America: systematic review on prevalence and variants. Mem Inst Oswaldo Cruz. 2014;109:553–68. doi:10.1590/0074-0276140123.

29. Whittaker C, Walker PGT, Alhaffar M, Hamlet A, Djaafara BA, Ghani A, et al. Under-reporting of deaths limits our understanding of true burden of covid-19. BMJ. 2021;375:n2239. doi:10.1136/bmj.n2239.

30. Andina Peruvian News Agency. Covid-19: segundo informe para actualizar cifra de fallecidos se conocerá esta semana. 26.07.2020. https://andina.pe/agencia/noticia-covid19-segundo-informe-para-actualizar-cifra-fallecidos-se-conocera-esta-semana-807333.aspx. Accessed 4 Sep 2025.

31. Sempé L, Lloyd-Sherlock P, Martínez R, Ebrahim S, McKee M, Acosta E. Estimation of all-cause excess mortality by age-specific mortality patterns for countries with incomplete vital statistics: a population-based study of the case of Peru during the first wave of the COVID-19 pandemic. Lancet Reg Health Am. 2021;2:None. doi:10.1016/j.lana.2021.100039.

32. Kupek E. How many more? Under-reporting of the COVID-19 deaths in Brazil in 2020. Trop Med Int Health. 2021;26:1019–28. doi:10.1111/tmi.13628.

33. Ministerio de Salud (MINSA). SINADEF - Defunciones Registradas. 2025. https://www.minsa.gob.pe/reunis/index.asp?op=1&niv=1&tbl=1#. Accessed 14 Aug 2025.

34. Castanheira HC, Da Monteiro Silva JHC. Examining sex differences in the completeness of Peruvian CRVS data and adult mortality estimates. Genus. 2022;78:3. doi:10.1186/s41118-021-00151-5.

35. Ministerio de Salud (MINSA). Fallecidos por COVID-19. 2025. https://www.datosabiertos.gob.pe/dataset/fallecidos-por-covid-19-ministerio-de-salud-minsa. Accessed 14 Aug 2025.

36. Ministerio de Salud (MINSA). Centro Nacional de Abastecimiento de Recursos Estratégicos en Salud (CENARES) - Distribucion por Producto. 2024. https://intranet.cenares.gob.pe/cenares/abastecimiento/DISTRIBUCION/wf_distribucion_por_producto.aspx. Accessed 5 Dec 2024.

37. Instituto Nacional de Salud (INS). Molecular Tests COVID-19: NETLAB. 2025. https://www.datosabiertos.gob.pe/dataset/dataset-de-pruebas-moleculares-del-instituto-nacional-de-salud-para-covid-19-ins. Accessed 11 Mar 2025.

38. Ministerio de Salud (MINSA). Non-Molecular Tests COVID-19: SISCOVID F100. 2025. https://www.datosabiertos.gob.pe/dataset/siscovid-f100-pruebas/resource/7f897963-f4c0-4a68-ae2f-10d49369edf9. Accessed 7 Feb 2025.

39. Ministerio de Salud (MINSA). Vacunación COVID19. 2025. https://www.datosabiertos.gob.pe/dataset/vacunacion/resource/c2296574-f1ca-46a4-9d1d-1aee320678a7. Accessed 20 Jun 2025.

40. Ministerio de Salud (MINSA). Población INEI 2021. 2025. https://datosabiertos.gob.pe/dataset/poblaci%C3%B3n-peru/resource/6e31247b-8057-450c-873d-d1ba28cf136a. Accessed 3 Aug 2025.

41. Ministerio de Salud (MINSA). Resolución Ministerial N.° 270-2020-MINSA; 2020.

42. Ministerio de Salud (MINSA). Resolución Ministerial N.° 375-2020-MINSA; 2020.

43. RCR Peru. CORONAVIRUS EN PIURA: EL 100% DE PACIENTES CON SÍNTOMAS LEVES SE HAN RECUPERADO CON IVERMECTINA. 2020. https://www.rcrperu.com/coronavirus-en-piura-el-100-de-pacientes-con-sintomas-leves-se-han-recuperado-con-ivermectina/. Accessed 4 Aug 2025.

44. Chamie JJ, Hibberd JA, Scheim DE. COVID-19 Excess Deaths in Peru’s 25 States in 2020: Nationwide Trends, Confounding Factors, and Correlations With the Extent of Ivermectin Treatment by State. Cureus. 2023;15:e43168. doi:10.7759/cureus.43168.

45. Soriano-Moreno DR, Fernandez-Guzman D, Sangster-Carrasco L, Quispe-Vicuña C, Grados-Espinoza P, Ccami-Bernal F, et al. Factors Associated With Drug Consumption Without Scientific Evidence in Patients With Mild COVID-19 in Peru. J Patient Saf. 2022;18:e1189–e1195. doi:10.1097/PTS.0000000000001053.

46. Quispe-Cañari JF, Fidel-Rosales E, Manrique D, Mascaró-Zan J, Huamán-Castillón KM, Chamorro-Espinoza SE, et al. Self-medication practices during the COVID-19 pandemic among the adult population in Peru: A cross-sectional survey. Saudi Pharm J. 2021;29:1–11. doi:10.1016/j.jsps.2020.12.001.

47. Castro NE, García DR, Rivera MT, Rondán-Guerrero P, García-Rojas F, Taype-Rondan A. Tendencias en el uso de fármacos para la COVID-19 durante la primera ola de la pandemia en un hospital de Lima, Perú. [Trends in drugs usage for COVID-19 during the first wave of the pandemic in a hospital in Lima, Peru]. Rev Peru Med Exp Salud Publica. 2021;38:608–14. doi:10.17843/rpmesp.2021.384.8820.

48. Casas Bueno, Guerrero Martínez. Medicamentos buscados en la web para la COVID-19 en Perú: Estudio infodemiológico [Tesis (Médico Cirujano)]. Lima, Perú: Universidad Peruana Cayetano Heredia; 2021.

49. Hoechle D. Robust Standard Errors for Panel Regressions with Cross-Sectional Dependence. The Stata Journal: Promoting communications on statistics and Stata. 2007;7:281–312. doi:10.1177/1536867X0700700301.

50. Fischer A, Michuda A. wildboottest (Python module); 2022.

51. Angulo-Bazan Y, Solis G, Cardenas F, Jorge A, Acosta J, Cabezas C. Household transmission in people infected with SARS-CoV-2 (COVID-19) in Metropolitan Lima; 2020.

52. Huamaní C, Velásquez L, Montes S, Mayanga-Herrera A, Bernabé-Ortiz A. SARS-CoV-2 seroprevalence in a high-altitude setting in Peru: adult population-based cross-sectional study. PeerJ. 2021;9:e12149. doi:10.7717/peerj.12149.

53. Moreira-Soto A, Pachamora Diaz JM, González-Auza L, Merino Merino XJ, Schwalb A, Drosten C, et al. High SARS-CoV-2 seroprevalence in rural Peru, 2021; a cross-sectional population-based study; 2021.

54. Moyano LM, Toledo AK, Chirinos J, Vilchez Barreto PMQ, Cavalcanti S, Gamboa R, et al. SARS-CoV-2 seroprevalence on the north coast of Peru: A cross-sectional study after the first wave. PLoS Negl Trop Dis. 2023;17:e0010794. doi:10.1371/journal.pntd.0010794.

55. Taype-Rondan A, Herrera-Añazco P, Málaga G. Sobre la escasa transparencia en los documentos técnicos para el tratamiento de pacientes con COVID-19 en Perú. Acta Med Peru 2020. doi:10.35663/amp.2020.372.982.

56. Caira-Chuquineyra B, Fernandez-Guzman D, Alvarez-Arias PM, Zarate-Curi ÁA, Herrera-Añazco P, Benites-Zapata VA. Association between prehospital medication and fatal outcomes in a cohort of hospitalized patients due to coronavirus disease-2019 in a referral hospital in Peru. Travel Med Infect Dis. 2022;50:102472. doi:10.1016/j.tmaid.2022.102472.

57. Martinez-Rivera RN, Taype-Rondan A. Overmedication in COVID-19 Context: A Report From Peru. J Clin Pharmacol. 2020;60:1155–6. doi:10.1002/jcph.1704.

58. Sindeev A, Martínez-Álvarez BM. Clinical and epidemiological characteristics of prisoners infected and deceased by COVID-19, National Penitentiary Institute of Peru, 2020. Rev Esp Sanid Penit. 2022;24:15–22. doi:10.18176/resp.00045.

